# Assessing the efficacy and safety of angiotensinogen inhibition using human genetics

**DOI:** 10.1101/2020.08.13.20174094

**Authors:** Jonas Bovijn, Jenny C Censin, Cecilia M Lindgren, Michael V Holmes

## Abstract

**Background:** Novel angiotensinogen (AGT) inhibitors are in early clinical development for treatment of hypertension. Evidence that this therapeutic approach will safely reduce risk of cardiovascular outcomes in humans is limited. We leveraged genetic data from more than one million individuals to characterise the effects of AGT inhibition.

**Methods:** We identified a genetic instrument for AGT inhibition from systolic blood pressure (SBP) genome-wide association study data, and investigated its relationship with *AGT* gene expression and circulating AGT protein concentration. We examined the instrument’s association with cardiovascular and renal outcomes, and compared the effect of the instrument with that of genetic instruments for other renin-angiotensin system (RAS) components and the causal effect of SBP overall. We performed phenome-wide association analyses to identify unanticipated effects of AGT inhibition.

**Results:** The AGT instrument (rs2478539; 0.49 mmHg lower SBP per G-allele) was strongly associated with hypertension, and showed evidence of colocalisation with *AGT* mRNA expression across various tissues. Scaled to a 10 mmHg lower SBP, the AGT instrument was associated with a 41% lower risk of major cardiovascular events, a composite of myocardial infarction, coronary revascularisation and stroke (111,549 cases; odds ratio 0.59, 95% confidence interval, 0.47 – 0.74; *P* = 3.1 × 10^-6^). There was little evidence of heterogeneity between the AGT vascular estimates when compared to equivalent estimates from other RAS targets and the effect of SBP lowering more broadly, and no strong evidence of potential target-mediated adverse effects.

**Conclusion:** Our findings suggest that inhibition of AGT safely reduces risk of major vascular events. These results support ongoing clinical development programmes for AGT inhibitors.

## Introduction

Hypertension is the leading contributor to global morbidity and mortality, with recent estimates indicating that elevated systolic blood pressure (SBP) leads to more than 200 million disability-adjusted life years and 10 million deaths annually.^1^ Lowering of blood pressure (BP) through lifestyle intervention or pharmacotherapy reduces the risk of several cardiovascular outcomes including coronary events and stroke.^2^ However, despite the existence of numerous BP-lowering pharmacotherapies, more than half of all patients with hypertension may have inadequately controlled BP^3^ (i.e. failing to lower their BP below the recommended threshold, typically 130/80 mmHg^4^). Several factors may contribute to this, including suboptimal adherence to therapy or lifestyle interventions, lack of access to healthcare, and physician prescribing behaviours, among others. Furthermore, resistant hypertension (the failure to control BP despite using a minimum of three antihypertensive drugs at maximal tolerated doses^5^) affects around 10% of patients with hypertension.^6^ These observations underscore a need for novel BP-lowering agents that are effective, safe and that promote greater adherence.

The renin-angiotensin system (RAS) is a hormone system that regulates blood pressure, primarily via its effects on fluid and electrolyte balance and systemic vascular resistance. Several proteins in the RAS pathway are targeted by commonly-used anti-hypertensive drugs (including angiotensin-converting enzyme (ACE) inhibitors, direct renin inhibitors, and angiotensin II receptor blockers^7^; Figure 1A). Angiotensinogen (AGT), a protein encoded by the *AGT* gene, is the initial substrate of this pathway, with cleavage of AGT by renin leading to production of angiotensin I. Angiotensin I is converted by ACE to angiotensin II, which is the primary mediator of the pathway’s effects on BP. At least two AGT-blocking biopharmaceutical agents are currently under development.^8^ These agents, an RNA interference (RNAi)-based therapy (ALN-AGT01) and an antisense oligonucleotide (ASO)-based therapy (IONIS-AGT-LRx), act by blocking translation of *AGT* messenger RNA (mRNA), thereby reducing the amount AGT protein produced. These therapeutic approaches allow for less frequent administration, which may offer benefits in terms of adherence and cardiovascular risk reduction, when compared to existing orally-administered small molecule-based antihypertensives.^8^ By way of example, recent clinical trials of inclisiran, an RNA-based inhibitor of proprotein convertase subtilisin/kexin type 9 (PCSK9), have shown that six monthly injection of this agent leads to sustained reductions in LDL cholesterol,^9^ which suggests that this approach may provide a means by which adherence is improved. Inhibition of AGT—the most proximal protein in the RAS pathway—may also avoid counterregulatory mechanisms that attenuate the BP-lowering effects of other RAS pathway targeting-drugs.^8^

**Figure 1.**
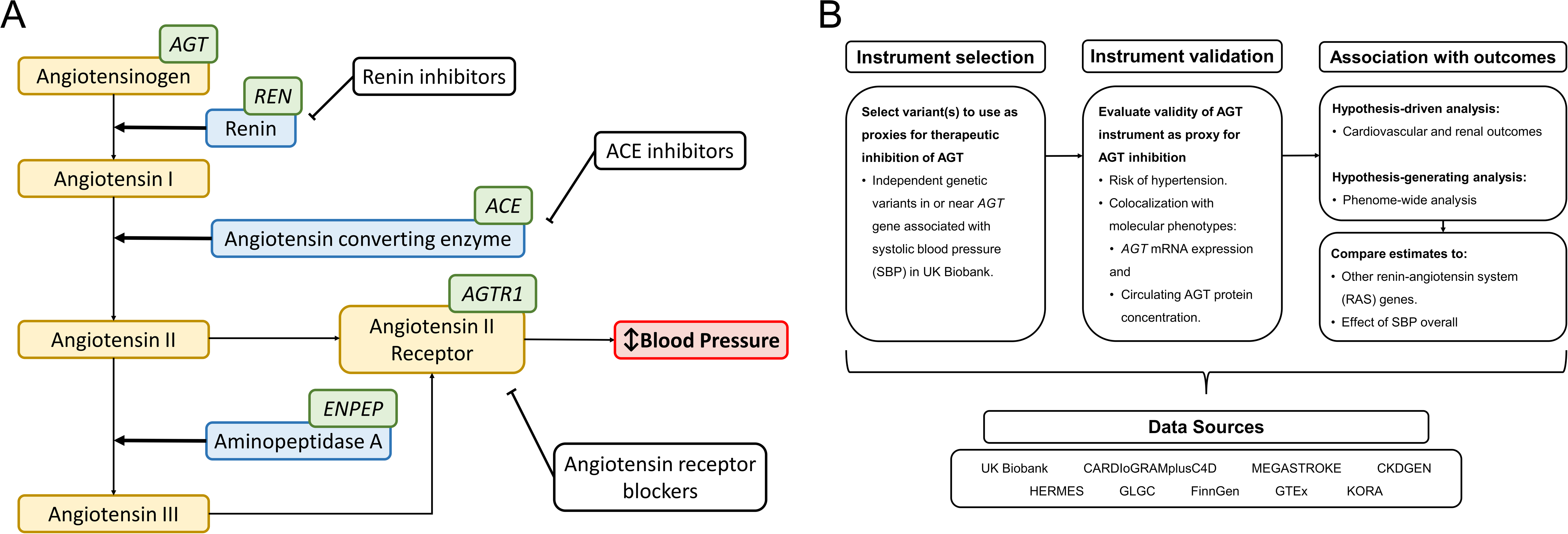
Renin-angiotensin system (RAS) and study design. **(A)** Simplified illustration of key components of the RAS. Yellow denotes main downstream mediators of AGT’s effect on blood pressure; blue denotes enzymatic proteins acting on the pathway; green denotes genes encoding various components of the pathway; existing RAS-targeting, anti-hypertensive drug classes are shown in white. Components not shown include *ACE2, AGTR2*, and others. **(B)** We selected genetic variants associated with systolic blood pressure (SBP) in or near to the *AGT* gene as proxies for therapeutic inhibition of AGT. We examined the validity of this instrument as a proxy for AGT inhibition by evaluating the instrument’s association with risk of hypertension and the likelihood of colocalisation with molecular phenotypes relating to *AGT*. The instrument’s association with clinical outcomes was examined in two stages. First, a hypothesis-driven analysis (focusing on cardiovascular and renal outcomes), and second, a hypothesis-generating analysis (encompassing phenome-wide analyses of clinical outcomes and biomarkers). Estimates relating to AGT were compared against those from other RAS genes and to those from an instrument encompassing all SBP-associated variants (i.e. genome-wide but excluding variants in or close to RAS pathway genes). CARDIoGRAMplusC4D, Coronary Artery Disease Genome wide Replication and Meta-analysis (CARDIoGRAM) plus The Coronary Artery Disease (C4D) Genetics; CKDGEN, Chronic Kidney Disease Genetics; GLGC, Global Lipid Genetics Consortium; GTEx, Genotype-Tissue Expression project; HERMES, Heart Failure Molecular Epidemiology for Therapeutic Targets; KORA, Cooperative Health Research in the Region of Augsburg.

Available pre-clinical evidence suggests that RNA-based AGT inhibitors lower BP in rodents^10,11^ and ALN- AGT01 and IONIS-AGT-LRx are currently under investigation in phase I and II randomised controlled trials (RCTs), respectively.^12-14^ Current guidance from the United States Food and Drug Administration (U.S. FDA) may enable anti-hypertensive approval on the basis of surrogate endpoints such as BP lowering,^15,16^ and allows for drug labelling that includes information on the cardiovascular risk reduction expected from such blood pressure lowering.^17^ For instance, aliskiren, a first-in-class small molecule direct renin inhibitor, was approved by the FDA in 2007 for the treatment of hypertension, without cardiovascular outcomes data at the time of approval.^18,19^ However, not all anti-hypertensive classes may offer similar reductions in cardiovascular and renal risk,^15,20-23^ and class- or drug-specific adverse effects may exist. Definitive conclusions about the efficacy and safety of AGT inhibition in humans may therefore require larger trials.

Naturally-occurring human genetic variation can be leveraged to validate the effects of drug target modulation in humans, and has been applied to a wide range of drug targets.^24-27^ Such studies may be particularly valuable prior to initiating large trials aimed at establishing effects on outcomes of interest. We applied this genetic approach to characterise target-mediated therapeutic and adverse effects that might be expected to arise from pharmacological inhibition of AGT.

## Methods

### Study design

We selected genetic variant(s) in or near to the *AGT* gene associated with SBP as proxies for AGT inhibition (Figure 1B). We examined the validity of this instrument as a proxy for AGT inhibition by evaluating the instrument’s association with risk of hypertension (using various phenotype definitions independent of reported BP values) and the likelihood of colocalisation with molecular phenotypes relating to AGT (AGT gene expression and circulating AGT protein concentration). The instrument’s association with clinical outcomes was examined in two ways, i.e. a hypothesis-driven analysis (focusing on cardiovascular and renal outcomes recognised to be associated with hypertension^28^) and a hypothesis-generating analysis (encompassing a phenome-wide analysis of binary outcomes and biomarkers). AGT estimates were compared to those from other RAS genes and to estimates from an instrument encompassing all SBP-associated variants (i.e. genome-wide but excluding variants in or close to RAS pathway genes).

### Renin-angiotensin system (RAS) pathway gene selection

We identified genes encoding well-established components of the RAS pathway, focusing on the most proximal components to AGT. This included *AGT* (encoding angiotensinogen), *REN* (renin), *ACE* (angiotensin-converting enzyme), *AGTR1* (angiotensin II receptor 1), and *ENPEP* (glutamyl aminopeptidase or aminopeptidase A). See Figure 1A for an illustrative diagram of the RAS pathway and its components. We excluded genes encoded on sex chromosomes *(AGTR2, ACE2)* due to a lack of sufficient genome-wide association study (GWAS) data pertaining to these genes.

### Instrument selection

We applied a stepwise conditional analysis algorithm^29^ to identify conditionally independent, GWAS-significant (*P* < 5 × 10^-8^) variants in or near (100 Kb on either side of each gene, with gene boundaries identified using the *biomaRt* R library) each target gene of interest, in publicly-available summary statistics from a GWAS of SBP.^30^ This dataset was selected since it was the largest set of publicly-available genome-wide summary statistics available for SBP in European-ancestry individuals that had not been adjusted for body mass index (which may induce issues related to collider or selection bias^31^) but which did include correction for medication use (addition of 15 mmHg to SBP for individuals reported to be taking BP-lowering medication), as is typically done for large-scale genetic association studies of blood pressure traits.^32^ Correcting for medication use in GWAS of SBP was recently shown not to lead to bias in Mendelian randomisation (MR) analyses of SBP.^33^

We examined each selected variant in the OpenTargets Genetics^34^ database to provide further evidence linking the variant to its intended target gene. This included annotations pertaining to the variant’s likely functional impact, distance between the variant and the target gene, and an aggregated score quantifying the functional evidence linking a variant to the target gene. If one of the selected variants was not available in one of the GWAS consortia datasets, we selected the best possible proxy (r^2^ > 0.9 in the CEU population, using the LDproxy function in the *LDLinkeR^35^* R library) to use instead.

### Data Sources and Outcomes

Estimates for the hypothesis-driven analysis were derived in UK Biobank (UKBB) and from publicly-available GWAS summary statistics for the relevant traits. The hypothesis-generating phenome-wide analyses were performed using data from UKBB and FinnGen. FinnGen is a project combining genotype data from Finnish biobanks and digital health record data from Finnish health registries.^36^

### UKBB: Population and Genotyping

UKBB is a prospective study of more than 500,000 British individuals aged between 45 and 69 and recruited between 2006 to 2010.^37^ Phenotypic data includes self-reported medical history at enrolment, as ascertained by self-administered touchscreen interface and a subsequent verbal interview with a medical professional; hospital-derived electronic health record (EHR) data, including International Classification of Diseases, ninth and tenth revision (ICD-9 and ICD-10) codes and Office of Population and Censuses Surveys Classification of Interventions and Procedures version 4 (OPCS-4) procedure codes, and an extensive set of physical measurements.

We excluded all samples indicated to have poor quality genotypes by UKBB (on the basis of high sample heterozygosity and missingness), and further excluded individuals meeting the following criteria: withdrawn their consent for participation; >10 third degree relatives; putative sex chromosome aneuploidy; sex mismatches (comparing genetically determined vs. self-reported sex, and comparing between assessments); ethnicity mismatches (mismatches between genetically determined and self-reported ethnicity for white British individuals, and any ethnicity mismatches between assessments). We reviewed pairwise genetic relatedness between individuals and excluded one individual per pair of individuals with an estimated second degree or closer relatedness (equivalent to a kinship coefficient of greater than 0.088). We included only individuals in the white British ancestry subset^38^ (i.e. samples who self-reported 'White British' and who have very similar genetic ancestry based on a principal components analysis of the genotypes). After applying these filters, up to 377,220 subjects remained.

Genotyping, quality control and imputation were performed centrally by UKBB, and details are fully described elsewhere.^38^ Briefly, genotype data are available for 488,377 individuals, 49,950 of whom were genotyped using the Applied Biosystems UK BiLEVE Axiom Array by Affymetrix [containing 807,411 markers^39^]. The remaining 438,427 individuals were genotyped using the Applied Biosystems UK Biobank Axiom Array by Affymetrix (containing 825,927 markers). Both arrays were specifically designed for use in the UKBB project and share ~95% of marker content. Phasing was done using *SHAPEIT3*, and imputation was conducted using *IMPUTE4*. For imputation, the Haplotype Reference Consortium (HRC) panel^40^ was used wherever possible, and for variants not in that reference panel, a merged UK10K + 1000 Genomes reference panel was used. SNPs were imputed from both panels, but the HRC imputation was preferentially used for SNPs present in both panels.

The UKBB project was approved by the North West Multi-Centre Research Ethics Committee and all participants provided written informed consent to participate. This research has been conducted under UKBB application number 11867.

### UKBB: Outcomes

Individual values for SBP in mmHg units were derived as in a recent GWAS for SBP performed in UKBB.^32^ We derived the mean SBP for each participant from two SBP measurements performed at baseline using either an automated or a manual sphygmomanometer. For subjects with only a single measurement available, we used this single value. We then corrected these values for blood pressure-lowering medication use by addition of 15 mmHg to the derived SBP value for subjects who had reported taking blood pressure medication at baseline.^41^ We defined hypertension in one of three ways, using either self-reported and/or physician-coded data (Table S1). We selected major cardiovascular events (MCVE; a composite of myocardial infarction [MI], coronary revascularisation and all stroke) as our primary outcome of interest, analogous to the commonly used composite outcome studied in cardiovascular outcomes trials. We also examined more specific vascular outcomes (myocardial infarction, coronary revascularisation, all stroke, ischaemic stroke, haemorrhagic stroke), and further cardiac (heart failure) and renal (chronic kidney disease and urinary albumin-creatinine ratio) outcomes. Definitions used in UKBB are given in Table S2.

### GWAS consortia

We supplemented data from UKBB with summary-level data from several GWAS, including data for MI,^42^ stroke (including all stroke and ischaemic stroke),^43^ haemorrhagic stroke,^44^ heart failure,^45^ chronic kidney disease,^46^ and urinary albumin-creatinine ratio.^47,48^ GWAS data for serum triglyceride concentration (TG) were also examined,^49,50^ following a putative association identified in the phenome-wide biomarker analysis. Where available, we selected data pertaining to analyses conducted in European-ancestry individuals. Further details on each consortium are provided in Table S3.

### Finngen

FinnGen is a public-private partnership project combining genotype data from Finnish biobanks and digital health record data from Finnish health registries, with summary statistic data currently publicly available for up to 96,499 participants (round 2 data release).^36^ A full description of the methods used to derive these data is given on the FinnGen web portal.^36^

## Statistical analyses

### Colocalisation analysis

We used *coloc*, a Bayesian modelling approach implemented as an R package^51^, to estimate the posterior probability of shared causal variant(s) driving associations in pairs of traits (i.e. colocalisation). We assessed colocalisation at the *AGT* locus for 2 pairs of traits: SBP and tissue-specific *AGT* mRNA expression (i.e. expression quantitative trait loci [eQTL] data),^52^ and SBP and circulating AGT protein concentration (i.e. protein quantitative trait loci [pQTL] data).^53^ We first ran colocalisation analyses using unconditioned summary statistics for each trait. Since the results from colocalisation analyses may be biased by the presence of multiple independent signals, we also ran colocalisation with conditioned datasets. To do this, we first identified independent signals in the *AGT* locus for each trait, using the GCTA-COJO statistical suite^29^ with genotype data from UKBB as a linkage disequilibrium (LD) reference panel. We then conditioned each dataset on the identified independent signals (using the GCTA-COJO -- cojo-cond function), and performed colocalisation analyses using these conditioned datasets.

The pQTL dataset included the effect allele and minor allele frequency but did not indicate the minor allele; this precluded derivation of the effect allele frequency in this dataset, which is required for conditional analysis. However, we extracted data pertaining to the minor allele and minor allele frequency for the same variants from a similar (European) ancestry population (i.e. UKBB) to identify the minor allele. To do this, we first filtered on variants with MAF < 0.45 in both datasets, and then filtered on variants where the difference in MAF between the two datasets was < 0.03. We then assigned the minor allele in the pQTL dataset, derived the effect allele frequency from the given minor allele frequency, and ran conditional analyses on these data.

We performed all colocalisation analyses on 400-kb and 2-Mb regions centred on the lead SBP variant, and used the default *coloc* priors. Colocalisation analyses where PP3 (the posterior probability of distinct causal variants) plus PP4 (the posterior probability of a shared causal variant) > 0.8 were considered to be adequately powered to detect colocalisation.^54^’^55^ For adequately powered colocalisation analyses, we considered a PP4 > 0.7 as being consistent with colocalisation between the two traits, similar to other recent studies.^56-59^ We visualised colocalisation using the *LocusCompareR* R library.^60^

### Association analysis in UKBB

The genotype-outcome association analyses in UKBB were performed using *SNPTEST* v2.5.4. We used an additive frequentist model (using *“-frequentist 1’)* and included sex, age at baseline, age^2^, genotyping array (a binary variable), recruitment centre, and the first 15 principal components as covariates in all analyses. We accounted for genotype uncertainty by using *“-method expected"*.

### Scaling and meta-analysis of estimates

We scaled all estimates relating to outcomes and traits of interest to a 10 mmHg reduction in SBP. To do this, we first derived the allelic effect estimate (in mmHg units) for the association of each instrument with SBP, by multiplying the effect estimate (in SD units, as per the selected GWAS in UKBB^30^) by the SD of medication-corrected SBP in UKBB (20.6 mmHg). We then derived a scaling factor for each instrument by dividing 10 by the allelic effect estimate for SBP and subsequently multiplied all per-allele effect estimates [log(OR) and the standard error of log(OR) for binary outcomes; beta and the standard error of beta for quantitative traits] by this scaling factor. Scaling may enable more meaningful interpretation of findings (as common genetic variants typically have small per-allele effect sizes) and allows for comparison between estimates from different instruments. However, genetic instruments for exposures represent potentially lifelong durations of exposure and the nature of the exposure-outcome relationship (for instance, whether it is cumulative) makes direct comparisons of estimates derived from a genetic instrument to a treatment trial challenging.

We meta-analysed scaled estimates from UKBB with scaled estimates from GWAS consortia for equivalent outcomes using inverse-variance weighted fixed-effect meta-analysis, implemented in the *metafor* R library.^61^

### Mendelian randomisation analysis of SBP

To derive MR estimates for the effect of genetically-predicted SBP on cardiovascular traits (MCVE, MI, all stroke and TG), we first extracted independent variants from a GWAS of SBP conducted in UKBB^30^ (the same dataset used for identifying the gene-specific instruments), using the *clump_data* function (with default settings of r^2^ cutoff < 0.001 and clumping window of 10,000 kb) in the *TwoSampleMR* R library.^62^ We removed all variants correlated (within 10 Mb and r^2^ > 0.001) with any of the RAS pathway genes’ instruments, which led to identification of 367 SBP-associated variants. Proxies (r^2^ > 0.9) were used if any variants were not present in the outcome datasets. We used the maximum number of SBP-associated SNPs available for each outcome; variants and estimates used in MR analyses are presented in Tables S4-S6. We then performed two-sample MR analyses of SBP on these outcomes, using summary statistics pertaining to studies with non-overlapping (with UKBB) participants (notably datasets from CARDIoGRAMplusC4D, MEGASTROKE, and Global Lipids Genetics Consortium). This included the inverse variance weighted-method, as well as the simple median, weighted median, and MR-Egger methods. The inverse variance weighted estimates were used if the estimates derived from the MR-Egger method suggested that there was no strong evidence of directional pleiotropy. MR estimates [log(OR) of outcome per SD unit change in SBP] were scaled to a 10 mmHg lower SBP by using the SD of medication-corrected SBP in UKBB (20.6 mmHg) and an MR estimate for MCVE was generated using fixed effect meta-analysis of the scaled MI and all stroke MR estimates. MR analyses were conducted using the *MendelianRandomization* R package.^63^

### Comparison to RAS pathway genes and SBP-instrument

We examined heterogeneity between the RAS (i.e. between *AGT, REN, ACE, and ENPEP)* estimates using Cochran’s Q test. We also compared the AGT estimates to those from the SBP MR analyses. Since the SBP MR analyses were performed using non-UKBB outcome data only (to avoid overfitting), we performed a sensitivity analysis where the AGT and RAS estimates only included data from the same datasets as used in the SBP MR analyses. We also performed these analyses for any putative associations found in the exploratory phenome-wide analyses (see below) to examine the potential validity of such associations and to assess whether the associations could represent target-specific effects as opposed to effects mediated through RAS or SBP. Lastly, we assessed the log-linear association between genetically lowered SBP and risk of vascular outcomes, comparing the effect of SBP overall (using the IVW estimates from the MR analyses of SBP and vascular outcomes, with the SBP instrument not depleted for RAS genetic variants) to the RAS gene-specific estimates. We performed phenome-wide scans in two large-scale cohorts: UKBB and the FinnGen cohort. For UKBB, we extracted estimates for the AGT instrument’s association with 1,074 binary outcomes (those with at least 200 cases available) from publicly available summary statistics generated by the Lee lab^64^ and for 30 circulating biomarkers analysed by the Neale lab.^65^ We used a 5% false discovery rate (FDR)-adjusted P-value threshold in each dataset. For FinnGen, we extracted estimates for 950 binary outcomes (those with at least 200 cases available) across 15 disease and organ-system categories, from publicly available summary statistics derived from up to 96,499 individuals,^36^ and also applied a 5% FDR-adjusted P-value threshold.

### Estimating the effect of therapeutic AGT inhibition

We sought to estimate the potential effect of therapeutic angiotensinogen inhibition on risk of vascular outcomes. One way of deriving such estimates is by comparing the effect of an instrument of interest (e.g. for AGT) to the effect of a reference instrument acting as a proxy for another, related, therapy that has already been evaluated in RCTs. Alternatively, if the effect of a drug target on disease appears to show a similar magnitude of effect as the downstream biomarker on which it intervenes (in this case SBP), it is possible to use a scaling factor derived from a polygenic SBP instrument as compared to estimates from meta-analysis of broader SBP-lowering agents. We used two reference instruments for this analysis: firstly, using genetic and therapeutic estimates for the overall effect of lowering SBP, and secondly, using genetic and therapeutic estimates for ACE inhibition. For the first analysis, we extracted estimates for the effect of pharmacologically lowered SBP on vascular outcomes from a meta-analysis of RCTs.^66^ To derive estimates pertaining to therapeutic AGT inhibition, we followed two steps. First, we determined a scaling factor by dividing the log(relative risk) of therapeutic SBP lowering by the log(OR) of genetically lowered SBP (with the genetic estimate derived using a ‘complete’ polygenic SBP instrument that was not depleted for RAS genetic variants). We then scaled the genetic AGT estimate by this scaling factor to estimate the pharmacologic AGT inhibition estimate per 10 mmHg lower SBP. For the second analysis, we extracted RCT estimates pertaining to ACE inhibitors from a Cochrane Systematic Review of first-line anti-hypertensive therapy.^67^ We extracted these estimates and first scaled them to a 10 mmHg reduction in SBP (using the effect of ACE inhibitors—a 16.5 mmHg reduction in SBP—as per the same meta-analysis^67^). To derive estimates pertaining to therapeutic AGT inhibition using the ACE-derived scaling factor, we followed the same steps outlined above.

## Results

### Selection and validation of AGT instrument

We identified one independent signal in the *AGT* gene associated with SBP at genome-wide significance (*P* < 5 × 10^-8^; rs2478539, 0.49 mmHg lower SBP per G-allele, *P* = 8.0 × 10^-27^; Figure S1). This variant, an intronic single nucleotide polymorphism (SNP) in *AGT*, was selected as a genetic proxy (i.e. instrument) for AGT inhibition. The instrument was strongly associated with risk of hypertension in UKBB, with consistent estimates for various definitions of this phenotype (e.g. clinician-coded hypertension, odds ratio [OR] 0.96 per G-allele, 95% confidence interval [CI], 0.95 – 0.97, *P* = 1.9 × 10^-9^, Figure 2A).

**Figure 2.**
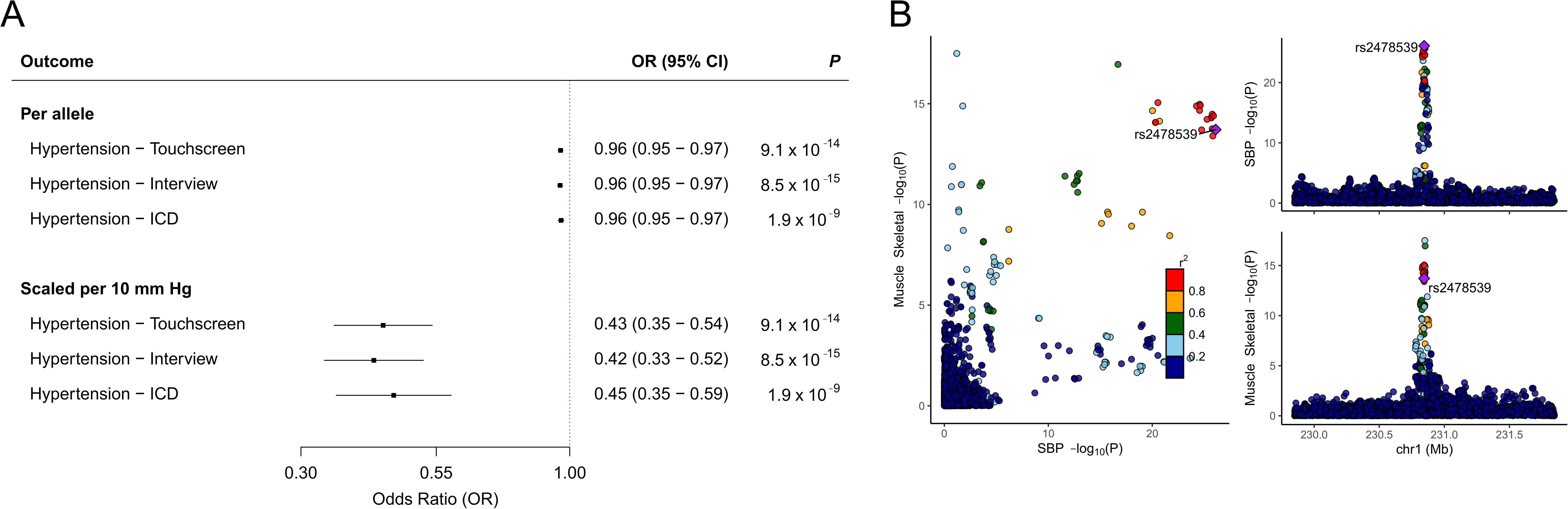
Validation of AGT instrument. **(A)** Per-allele association and scaled estimates of the AGT instrument (comprising rs2478539) with risk of hypertension in UK Biobank (UKBB), using various definitions of hypertension. “Touchscreen” refers to participants self-reporting (at enrolment) on a touchscreen interface that a doctor has ever told them that they have high blood pressure; “Interview” refers to nurse-led interview at enrolment where participants reported having hypertension; “ICD” refers to International Classification of Disease version 9 and 10 codes [see methods for codes included in definition] for hypertension, as recorded by hospital clinicians in the course of routine clinical care in the National Health Service (NHS). Boxes represent point estimates of effects. Lines represent 95% confidence intervals (CI). OR, odds ratio. **(B)** Colocalisation of *AGT* mRNA expression (i.e. expression quantitative trait loci) in skeletal muscle tissue (bottom right frame) and SBP (top right frame). Each plotted point represents a SNP; colours of the plotted points indicate the linkage disequilibrium (in r^2^) of each SNP with rs2478539 (the main AGT instrument used in our analyses and indicated as a purple point). The posterior probability of a shared causal variant (PP4) in this analysis was 86%.

To further investigate the validity of the instrument as a proxy for investigational RNA-based AGT inhibitors, we used colocalisation analyses to examine the effect of this variant on *AGT* mRNA expression across several tissues and circulating AGT protein concentration. Colocalisation analyses of SBP and *AGT* mRNA expression (using expression quantitative trait loci [eQTL] data from several tissues^52^) provided evidence of colocalisation in several tissues, using both unconditioned datasets and datasets conditioned on either the lead eQTL or the lead SBP-associated variant (e.g. probability of a shared causal variant [PP4] in skeletal muscle tissue = 86% in the unconditioned analysis; Figure 2B, Table S7-S8).

Next we evaluated the effect of the instrument on circulating AGT protein concentration,^53^ as recent evidence from a phase I RCT has shown that administration of ALN-AGT01 leads to large reductions in circulating AGT protein levels.^68^ We found a 0.13 standard deviation (SD) units lower AGT concentration (95% CI, -0.22 – -0.05); *P* = 0.003; Figure S2) per SBP-lowering allele of rs2478539. There was no evidence of colocalisation of genetic signals relating to AGT plasma concentration (i.e. protein quantitative trait loci [pQTLs]) and the lead SBP signal (PP4 < 1%; Figure S3A, Table S9); however, using data conditioned on the lead SBP signal did provide evidence of colocalisation (PP4 = 88%; Figure S3B, Table S9). We selected the top AGT pQTL (rs73102646, r^2^ = 0.21 and D’ = 0.94 with rs2478539 in European-ancestry populations) for sensitivity analysis.

### Association with cardiovascular and renal outcomes

We investigated the association of the AGT instrument with cardiovascular outcomes, including coronary events (MI and coronary revascularisation), stroke (haemorrhagic and ischaemic stroke) and major cardiovascular events (MCVE; a composite of all coronary events and strokes, including death from either), scaled to a 10 mmHg lower SBP. This revealed a 41% lower risk of MCVE (OR 0.59; 95% CI, 0.47 – 0.74; *P* = 3.1 × 10-^6^), 45% lower risk of MI (OR 0.55; 95% CI, 0.40 – 0.76; *P* = 2.6 × 10^-4^), and a 31% lower risk of all stroke (OR 0.69; 95% CI, 0.50 – 0.94; *P* = 0.02), with similar reductions in risk of coronary revascularisation and haemorrhagic stroke (Figure 3; Figure S4). Estimates for MCVE, MI and all stroke derived using the AGT pQTL were concordant to those of our primary AGT instrument, scaled to the same difference in SBP (P-heterogeneity = 0.82, 0.98, and 0.45 respectively for MCVE, myocardial infarction and all stroke, with similar findings when comparing conditional estimates; Figures S5-S6).

**Figure 3.**
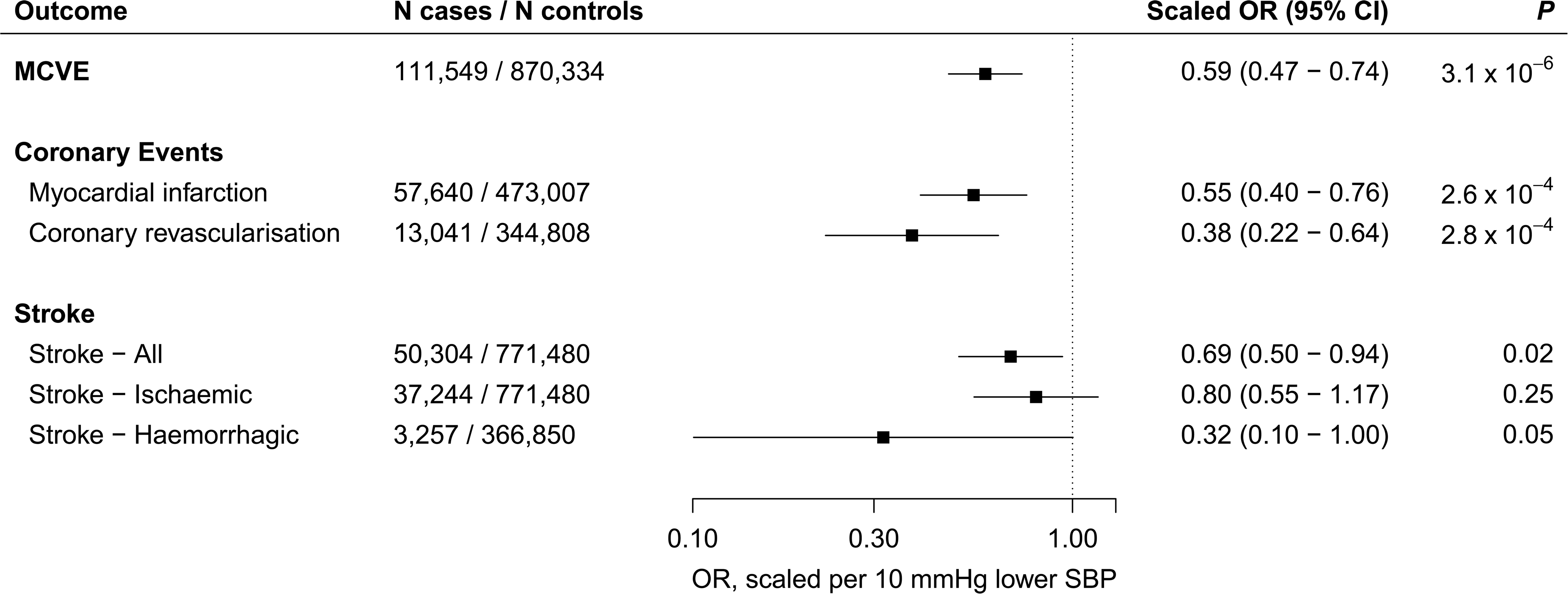
Scaled associations of the AGT instrument with cardiovascular outcomes. Scaled estimates of the AGT instrument with risk of major cardiovascular events (MCVE; a composite of MI, coronary revascularisation and all stroke), MI, coronary revascularisation, all stroke, ischaemic stroke, and haemorrhagic stroke. Estimates are scaled to a 10 mmHg lower SBP and aligned to the blood pressure-lowering allele of rs2478539. Boxes represent point estimates of effects. Lines represent 95% confidence intervals. CI, confidence interval; OR, odds ratio.

The AGT instrument was weakly associated with a lower urinary albumin-creatinine ratio (-0.07 ln(mg/g) units, 95% CI, -0.14 – -0.01, *P* = 0.03; Figure S7) and lower risk of chronic kidney disease (OR 0.72; 95% CI, 0.51 – 1.01; *P* = 0.06; Figure S8). We found no evidence of association with risk of heart failure (OR 0.92; 95% CI, 0.67 – 1.27; *P* = 0.63; Figure S8). We assessed association with further outcomes with phenome-wide association analyses (PheWAS).

### Comparison to RAS pathway genes and a genome-wide SBP instrument

We next compared the effects of the AGT instrument to those of other downstream targets in the RAS pathway (Figure 1A). We followed an identical procedure to that used to select the AGT instrument, and identified instruments for three additional components of the RAS (renin, angiotensin-converting enzyme and aminopeptidase A, encoded by *REN, ACE*, and *ENPEP*, respectively; Table S10). We also compared the AGT estimates to those relating to an instrument including up to 364 SBP-associated variants (i.e. genome-wide but excluding variants in or close to the selected RAS genes; see Table S4-S5 for variants included in the SBP instrument and Tables S11-S12 for results of the SBP MR analyses). We found no strong evidence of heterogeneity between the RAS targets’ estimates for MCVE, MI or all stroke (Figure 4A-C). The AGT estimates were directionally concordant to the SBP instrument’s estimates for these three vascular outcomes, with some evidence of heterogeneity between estimates for MI and MCVE (Figure 4A- C). Sensitivity analysis which included restricting analyses to the same data sources (i.e. only including data from the MEGASTROKE and CARDIoGRAMplusC4D consortia) attenuated the heterogeneity between the AGT and SBP instruments’ estimates (Figure S9A-C). The per-allele effects of each of the RAS drug-target instruments (i.e. AGT, REN, ACE and ENPEP) demonstrated disease associations that were consistent with the overall log-linear relationship between SBP and vascular outcomes (Figure 4D-F).

**Figure 4.**
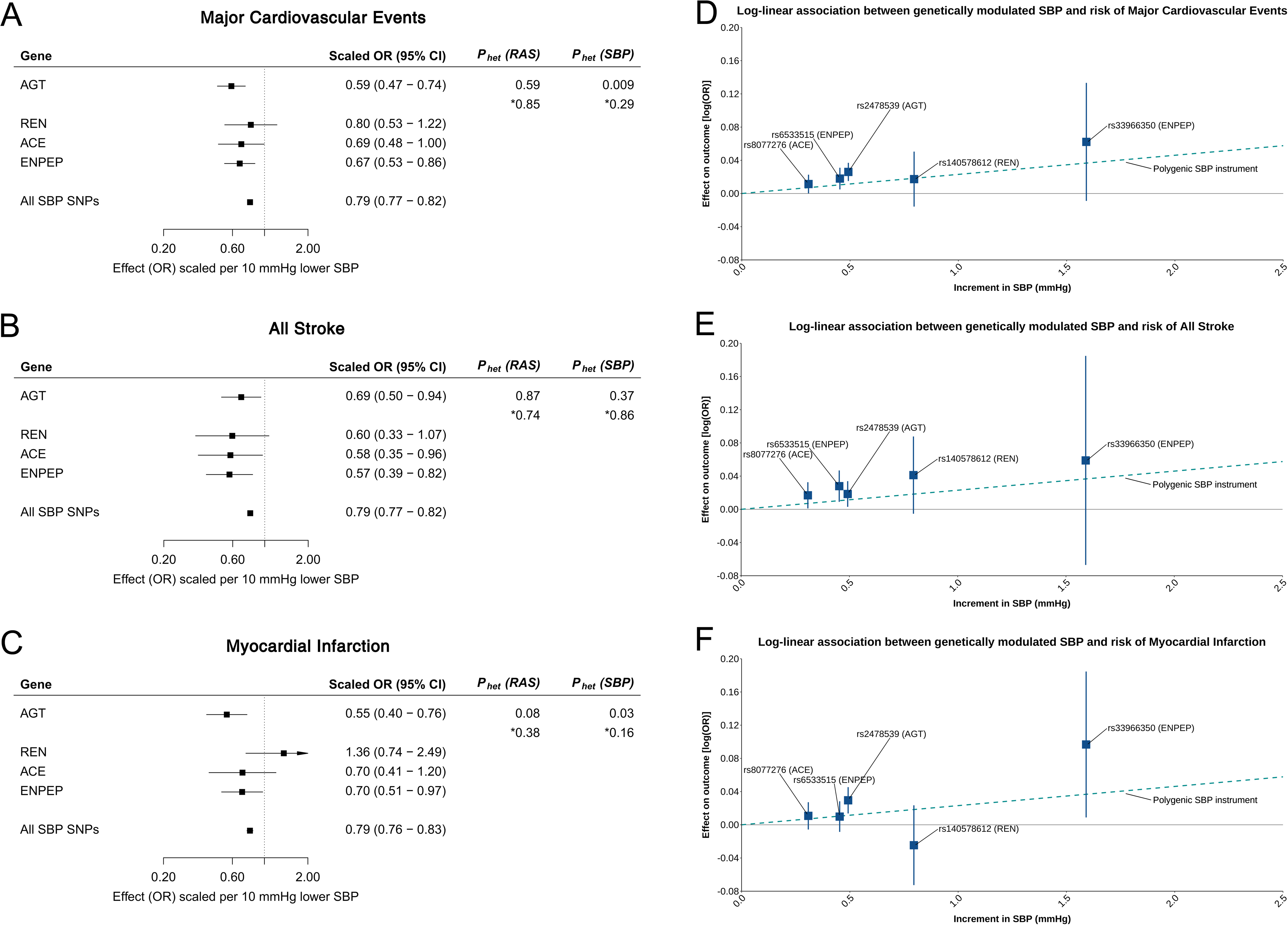
Comparison of scaled associations of AGT, RAS pathway genes and a genome-wide SBP-instrument and log-linear associations between genetically modulated SBP and risk of vascular outcomes. The left-hand panels (A-C) present the scaled estimates of SBP-lowering instruments for *AGT* (rs2478539), *REN* (rs140578612), *ACE* (rs8077276), *ENPEP* (rs6533515 and rs33966350) and an “all SBP-SNPs” instrument (up to 364 SBP-associated variants and excluding any variants correlated with RAS pathway gene variants) with risk of (A) major cardiovascular events (MCVE), (B) all stroke, and (C) MI. Estimates are scaled to a 10 mmHg lower SBP. P_het_(RAS) refers to P-value from Cochran’s Q test for heterogeneity between the RAS pathway estimates (i.e. between AGT, REN, ACE, and ENPEP estimates). P_het_(SBP) refers to P-value from Cochran’s Q test for heterogeneity between the AGT estimate and the SBP-instrument estimate. Values marked with “*” refer to heterogeneity estimates derived in sensitivity analysis restricted to the same datasets, to enable reliable comparisons that are not influenced by between-study heterogeneity (using non-UKBB data only, see Figure S9 for further information). The right-hand panels (D-F) present the per-allele effect [log(OR)] of each RAS pathway instrument (blue boxes) and the overall effect of genetically lowered SBP (dotted lines, representing the effect of all SBP-associated variants, including any variants correlated with RAS pathway gene variants), on (D) major cardiovascular events (MCVE), (E) all stroke, and (F) MI. Boxes represent point estimates of effects. Lines represent 95% confidence intervals (CI). OR, odds ratio; SNPs, single-nucleotide polymorphisms.

### Association with further outcomes

A PheWAS of 1,074 binary outcomes in UKBB revealed no strong evidence of further associations at a 5% false-discovery rate (FDR) threshold (Figure S10). PheWAS of 950 outcomes in up to 96,499 Finnish individuals revealed a top association of “Hypertensive diseases” (supporting the validity of the instrument in this cohort), and no further association at the same FDR threshold (Table S13). Investigation of 30 circulating biomarkers in UKBB revealed an association of the AGT instrument with elevated plasma triglyceride concentration (TG; 0.16 SD units higher scaled to a 10 mmHg lower SBP, 95% CI, 0.07 – 0.26, *P* = 8.2 × 10^-4^, *P*_FDR-adjusted_ = 0.02; Table S14). There was little evidence of association with TG in an independent GWAS of blood lipids^49^ (0.07 SD units higher, *P* = 0.34; Figure S11). There was also evidence of heterogeneity between the AGT TG estimate and estimates relating to the RAS pathway genes and the SBP instrument, in contrast to the relative concordance between the vascular estimates (Figure S12; see Table S6 for variants included in the SBP instrument and Table S15 for results of the SBP MR analysis). A further colocalisation analysis revealed no strong evidence of colocalisation between genetic signals pertaining to SBP and TG at the *AGT* locus (probability of distinct causal variants [PP3] = 73% and probability of shared causal variant [PP4] = 4%, using a 2Mb-interval; Figure S13, Table S16).

### Estimating the expected clinical effect of pharmacological AGT inhibition

Estimates derived from human genetics may differ to those from clinical trials for multiple reasons, including that the effects of genetic variants may be lifelong, as opposed to pharmacological perturbation of shorter duration and occurring later in life.^69^ To gauge insight into the expected effect size of pharmacological inhibition, we can scale the genetic estimates for a target of interest by a scaling factor derived from an existing biomarker or drug target for which there are RCT data and a valid genetic instrument, and where the mechanism of action is related to that of our target of interest.^69^ Figure 4 shows that the vascular disease associations for genetic instruments for individual RAS therapeutic targets scale to the overall association of a polygenic instrument for SBP. This suggests that a scaling factor derived from SBP (comparing a polygenic SBP genetic instrument to meta-analysis of SBP lowering agents) may be a reliable approach to estimating the therapeutic effect of AGT inhibition. When we compared an SBP genetic risk score to meta-analysis of SBP lowering agents,^66^ we found very similar magnitudes of disease association (for MCVE this was OR 0.79; 95% CI 0.77 – 0.82 for the SBP genetic instrument and RR 0.80; 95% CI 0.77 – 0.83 for meta-analysis of RCTs; both per 10mmHg lower SBP). The corresponding values for genetic and pharmacological inhibition of ACE were OR 0.69; 95% CI 0.48 – 1.00 for the ACE genetic instrument and RR 0.85; 95% CI 0.79 – 0.91 for meta-analysis of RCTs^67^ (Figure 5). Applying these scaling values to the genetic data for AGT, we estimate that an RCT that lowers SBP by 10mmHg will achieve ORs of MCVE of between 0.60 to 0.79 (point estimates scaled using SBP-derived and ACE-derived transformations, respectively).

**Figure 5.**
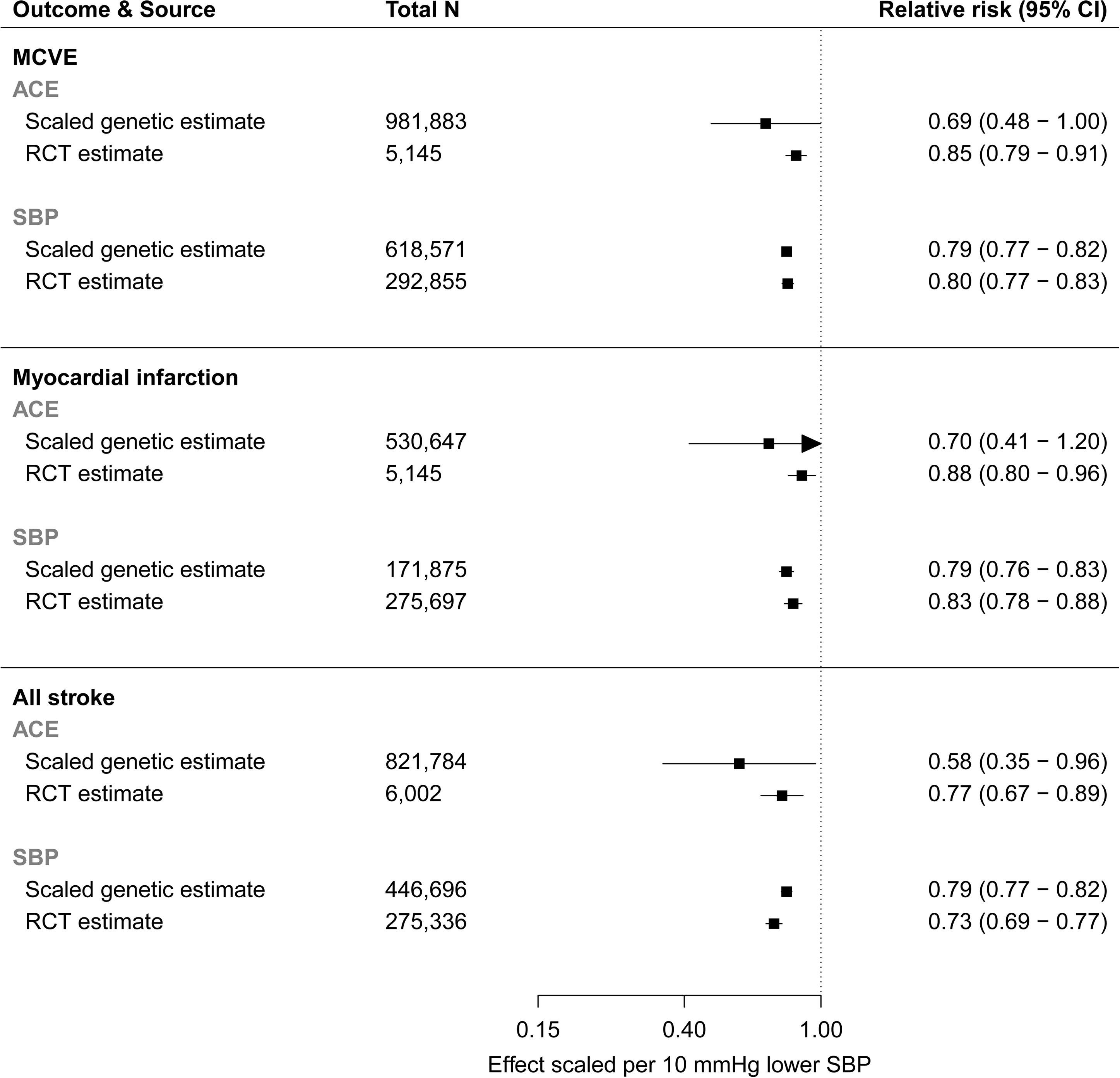
Triangulation of genetic and RCT estimates to predict estimates for therapeutic inhibition of AGT. RCT and genetic estimates relating to SBP lowering (up to 372 SBP-associated variants) and ACE inhibition (rs8077276) are shown. RCT estimates for SBP lowering were derived from a meta-analysis of SBP-lowering RCTs^66^ and RCT estimates for ACE inhibition were derived from a Cochrane systematic review of first-line anti-hypertensive therapy.^67^ All genetic and RCT estimates were scaled to a 10 mmHg lower SBP. Lines represent 95% confidence intervals. RCT, randomised controlled trial.

## Discussion

We leveraged large-scale human genetic datasets to show that a genetic instrument for AGT inhibition is associated with lower risk of vascular events, with no strong evidence of major harmful associations. These findings suggest that therapeutic inhibition of AGT may be expected to safely reduce risk of vascular outcomes without leading to major target-mediated adverse effects.

While novel AGT inhibitors may potentially be granted a marketing authorisation on the basis of a surrogate outcome (i.e. their effect on BP), larger trials will be needed to establish whether these agents safely reduce cardiovascular risk. Available animal data support the efficacy and safety of AGT inhibition,^10,11^ but translation of evidence from animals to humans is recognised to present numerous challenges and limitations.^70,71^ Randomised studies in humans therefore provide more reliable evidence of efficacy and safety;^72^ human genetics provides one way of conducting such investigations. Furthermore, while existing RAS targeting medicines have generally been found to be safe, the possibility exists that inhibition of upstream targets in RAS (such as AGT) may lead to unique, non-RAS mediated effects not previously observed in studies of other RAS pathway modulators. Human genetic data also allows for exploration of such target-specific effects.^73^

The application of human genetics to investigating the effects of drug target modulation is now well-established,^73^ with recent examples including investigations of cardiovascular safety,^27^ identification of novel drug indications^74^ and discovery of opportunities for precision medicine.^75^ Previous studies of genetic proxies for glucagon-like peptide-1 receptor (GLP-1R) modulation^25^ and Niemann-Pick C1-like 1 (NPC1L1) inhibition,^76^ among others, have successfully anticipated cardiovascular risk reduction prior to completion of cardiovascular outcomes trials for these drug classes. Recent work used variants related to existing anti-hypertensive medications to recapitulate known therapeutic effects and discover novel potential adverse effects.^77^ Our study applied these approaches to provide several insights into the effects of AGT inhibition.

First, we find that genetic variation in *AGT* is strongly associated with lower risk of major vascular events, particularly coronary events, and nominally associated with lower risk of stroke. The AGT instrument’s association with lower albuminuria and reduced risk of chronic kidney disease suggests that AGT inhibition may also display beneficial effects on these outcomes, in line with the established therapeutic role of RAS pathway inhibition in these renal phenotypes.^78^ Numerous candidate-gene studies have studied the association of genetic variation in *AGT* with hypertension and vascular outcomes, with conflicting and inconsistent results.^79^ The reasons for these discrepancies are potentially manifold, and include limitations related to sample size, population structure, linkage disequilibrium, and others.^80^ Our study integrates data from large-scale GWAS, transcriptomic and proteomic investigations (including more than one million participants) to address such limitations and shows that variation in *AGT* is robustly associated with hypertension and vascular outcomes, and that these genetic associations are likely mediated by effects on *AGT* transcription. The proximity of this molecular mediator (gene expression) to the mechanism of RNA-based AGT inhibitors (blockade of mRNA translation) strengthens the relevance of our findings in the context of drug target validation.

Second, the concordance of the AGT vascular estimates to those relating to other RAS pathway genes suggests that these effects are RAS-pathway mediated, whilst the congruence to the estimates of the SBP-instrument further indicates that these effects are directly related to the RAS pathway’s BP-lowering effect. These findings indicate that the AGT instrument’s associations with these outcomes represent true, target-mediated effects.

Third, by comparing our genetic AGT estimates to genetic and RCT estimates relating to SBP lowering and ACE inhibition, we approximated the expected effect of therapeutic AGT inhibition on vascular outcomes. The AGT estimates derived in these two analyses may represent plausible ranges for the expected effect of therapeutic AGT inhibition on vascular outcomes, particularly as we did not observe strong evidence of heterogeneity between the AGT genetic estimates and those pertaining to the other RAS genes or the SBP-instrument. It is worth noting several potential caveats to such comparisons, which may include imprecision in estimates used to derive scaling factors (such as e.g. the ACE genetic estimates), AGT inhibitors and/or other SBP-lowering therapies having off-target effects, comparative trials not being of a similar duration, differential adherence, and dissimilar trial participant characteristics (including background therapy), among others.

Recent non-peer reviewed data from a phase I study^68^ suggest that ALN-AGT01 leads to a more than 90% reduction in AGT levels and a more than 10 mmHg reduction in mean 24-hour SBP at week 8, compared to placebo. IONIS-AGT-LRx is currently in phase II development^13^, with no reported phase I data to date. Our phenome-wide association analyses in two large-scale European cohorts found no strong evidence of target-mediated adverse effects, supporting a phase I report of ALN-AGT01 that has thus far noted no drug-related serious adverse events.^68^ Although these early results are encouraging, larger phase II and III studies will be needed to further verify our genetic findings and quantify an absence of off-target effects.^81^

Fourth, our work highlights several issues that are more broadly relevant to studies applying large-scale human genetic data to investigate drug targets.^82^ Genetic variants used as proxies or surrogates for drug target modulation are typically selected from genetic association data relating to either gene expression, protein concentration or downstream biomarkers. In the present study, we selected variants on the basis of their association with a clinical biomarker (i.e. SBP), similar to previous studies of other drug targets.^27,83,84^ Previous studies have linked genetic variation in *AGT* with altered *AGT* gene expression and/or AGT protein levels, though such findings were inconsistent, largely owing to the limitations of such candidate gene studies as mentioned above. We applied colocalisation to provide more definitive evidence linking SBP-associated variation in *AGT* to *AGT* transcription and/or translation. Colocalisation of SBP with *AGT* mRNA expression suggests that altered gene expression may be a mediating mechanism. We found that our AGT instrument was also associated with circulating AGT protein concentration (as measured using an aptamer-based assay^53^), although this association was not supported by colocalisation when using unconditioned estimates. Whilst this could represent a true finding (i.e. distinct genetic variants drive associations with SBP and AGT protein concentration), there may be alternative explanations. For instance, previous work has shown that the presence of protein-altering variants may lead to altered binding of the aptamer to the target protein, biasing measurement of protein abundance.^85,86^ The presence of a missense variant in *AGT* (rs699) in high LD with the selected AGT instrument (r^2^ = 1.0 in European populations) may influence aptamer binding, and could therefore attenuate the association of the AGT instrument with AGT protein concentration (Figure S14). Personal communication with the manufacturer of the SomaLogic platform confirmed that the location of the AGT aptamer-binding site is currently unknown (SomaLogic CO, USA; August 2019). Further work leveraging alternative technologies for measuring circulating protein levels (e.g. the antibody-based proximity extension assay offered by Olink Proteomics^87^) may yield more reliable results in this regard. Nevertheless, the similarity of the estimates relating to the pQTL-sourced and the SBP-sourced AGT instruments, and evidence of colocalisation of *AGT* mRNA expression with SBP support the validity of our instrument as a proxy for AGT inhibition.

Our analysis of circulating biomarkers identified a potential association with elevated TG, which, although modest in the context of the multiple phenotypes tested, prompted further investigation. Whilst this association may reflect a true causal relationship (i.e. genetic variants mimicking therapeutic inhibition of AGT cause higher TG, which implies that pharmacological inhibition may also be expected to lead to higher TG), several other explanations may be worth considering (Figure S15).^82^ The heterogeneity observed between the AGT TG estimate and the estimates relating to other RAS pathway genes, suggests that the association of the AGT instrument is unlikely to be mediated via the RAS pathway (in contrast to the concordant associations observed for vascular traits). As AGT lies upstream from other RAS targets, the potential arises for target-mediated yet non-RAS or non-SBP mediated effects ((B) in Figure S15),^73^ with the implication that, in this scenario, therapeutic AGT inhibition may still lead to a rise in TG through target-mediated mechanisms. Evidence in rodents suggests that AGT may have non-RAS mediated effects,^88^ however several further explanations may exist (Figure S15). The AGT variant’s association with TG may be biased by horizontal (also termed pre-translational^89^) pleiotropy or linkage disequilibrium, or be a chance (i.e. false-positive) finding. Our colocalisation analysis of TG and SBP at the *AGT* locus suggests that distinct causal variants are associated with these traits and that the AGT instrument’s association with TG may therefore be confounded by linkage disequilibrium. However, if the AGT instrument was in linkage disequilibrium with a TG-increasing variant, we would expect the AGT instrument’s association with cardiovascular outcomes to be similarly influenced. For instance, in a recent MR study investigating the causal effect of blood lipids on coronary disease^90^, a 1 SD higher TG was shown to lead to a 34% higher risk of coronary disease. Assuming the relationship between TG and vascular disease is linear, the AGT instrument’s effect on TG (0.14 SD higher) would therefore be expected to lead to an approximately 4% increase in risk of coronary disease. The AGT instrument’s strong association with lower risk of coronary events, similar to that expected on the basis of SBP lowering, suggests that this is unlikely, although our analyses may be underpowered to detect an effect of this magnitude. This, together with the lack of replication in an independent dataset,^49^ therefore suggests that the TG association found in UKBB is likely to be a chance finding. Nevertheless, whilst these investigations suggest that TG elevation is unlikely to be a target-mediated effect of AGT inhibition, monitoring of blood lipids in a subset of individuals enrolled in ongoing clinical trials of AGT inhibition may be potentially warranted.

Our study has several limitations. Firstly, we have shown evidence of colocalisation with *AGT* mRNA expression in various tissues, albeit not in liver tissue, which is the primary target tissue for RNA-based AGT inhibitors currently under development. Our analyses were underpowered to detect evidence of colocalisation in liver tissue and larger sample sizes may improve this (e.g. in GTEx, liver tissue only had 153 samples, vs. 491 samples for skeletal muscle, where we did find evidence of colocalisation). However, sensitivity analysis using a variant associated with circulating AGT protein concentration, which is mainly liver-derived,^8^ yielded similar estimates to that of our instrument. In light of this and recent data showing that ALN-AGT01 lowers circulating AGT levels^68^, our findings are likely to be of relevance to these novel therapies. Second, our methods would not be able to predict potential off-target effects of specific agents or modalities (such as, for instance, torcetrapib-mediated increases in BP, an adverse effect with BP increased to a greater extent than that observed for other CETP inhibitors^91^), nor are they likely to detect idiosyncratic adverse effects. Nonetheless, human genetics may be helpful in evaluating whether effects observed in clinical trials are target-mediated or not,^73^ as recently shown for volanesorsen-associated thrombocytopenia. Loss-of-function genetic variants in *APOC3* were shown not to be associated with platelet counts,^92^ which suggests that the thrombocytopenia observed in a prior trial of volanesorsen,^93^ an ASO targeting apolipoprotein C-III, was not target-mediated. Recent data announced for AKCEA-APOCIII-LRx, a newer-generation ASO also targeting apolipoprotein C-III, revealed no effects on platelet count.^94^ This suggests that the platelet-effect observed with volanesorsen was not target-mediated, and showcases the utility of human genetics in disentangling effects observed in clinical trials. Third, analyses may be underpowered to detect rare effects, even if such effects are target-mediated, particularly given the relatively small effect size of the instrument on the exposure. The phenome-wide analyses we have undertaken may therefore have missed uncommon effects. Finally, the effects relating to potentially life-long genetically modulated AGT may not be directly comparable to those of short-term pharmacological modulation in adulthood, although comparison to a reference standard^69^ (SBP lowering and ACE inhibition in our case) may yield more therapeutically-relevant estimates. Comparison of novel AGT inhibitors (RNA-based therapeutics) to existing anti-hypertensives (typically small molecules) may however still present challenges (for instance, improved adherence—a recognised independent predictor of cardiovascular risk^95^—with RNA-based therapeutics may lead to greater reductions in cardiovascular risk than observed with small molecules).

In conclusion, our findings provide evidence to support clinical trials of AGT inhibitors as likely showing these agents reduce risk of major cardiovascular events, with no strong evidence for target-mediated adverse effects. Our results illustrate the value of applying human genetics to shed light on effects of novel therapeutic targets, particularly in settings where such therapies may be approved on the basis of surrogate outcomes.

## Acknowledgments

We express our gratitude to the studies, consortia and biobanks included in this study, especially their investigators and their participants, for their valuable contributions to our research. We thank Dr A. Ndungu for advice pertaining to one of our analyses.

## Contributors

J.B., M.V.H. and C.M.L. contributed to the conception and design of the study. All authors contributed to the data collection and analyses and interpretation of findings. J.B., M.V.H. and C.M.L. contributed to the first draft of the manuscript. All authors contributed to and approved the final version of the manuscript. The corresponding authors had full access to all the data in the study and shared final responsibility for the decision to submit for publication with all authors.

## Data availability

All GWAS summary statistics used in our analyses are publicly available. Access to individual-level UKBB data is available on application to UKBB.

## References

1 Forouzanfar MH, Afshin A, Alexander LT, et al. Global, regional, and national comparative risk assessment of 79 behavioural, environmental and occupational, and metabolic risks or clusters of risks, 1990-2015: a systematic analysis for the Global Burden of Disease Study 2015. Lancet 2016; 388:1659–724.

2 Xie X, Atkins E, Lv J, et al. Effects of intensive blood pressure lowering on cardiovascular and renal outcomes: updated systematic review and meta-analysis. Lancet 2016; 387: 435–43.

3 Wang TJ, Vasan RS. Epidemiology of uncontrolled hypertension in the United States. Circulation 2005; 112: 1651–62.

4 Bakris G, Ali W, Parati G. ACC/AHA Versus ESC/ESH on Hypertension Guidelines: JACC Guideline Comparison. J Am Coll Cardiol 2019; 73: 3018–26.

5 Sarafidis PA, Georgianos P, Bakris GL. Resistant hypertension--its identification and epidemiology. Nat Rev Nephrol 2013; 9: 51–8.

6 Noubiap JJ, Nansseu JR, Nyaga UF, Sime PS, Francis I, Bigna JJ. Global prevalence of resistant hypertension: a meta-analysis of data from 3.2 million patients. Heart 2019; 105: 98–105.

7 Zaman MA, Oparil S, Calhoun DA. Drugs targeting the renin-angiotensin-aldosterone system. Nat Rev Drug Discov 2002; 1: 621–36.

8 Ren L, Colafella KMM, Bovée DM, Uijl E, Danser AHJ. Targeting angiotensinogen with RNA-based therapeutics. Curr Opin Nephrol Hypertens 2020; 29: 180–9.

9 Fernández-Ruiz I. Twice-yearly inclisiran injections halve LDL-cholesterol levels. Nat Rev Cardiol 2020; 17: 321.

10 Uijl E, Mirabito Colafella KM, Sun Y, et al. Strong and Sustained Antihypertensive Effect of Small Interfering RNA Targeting Liver Angiotensinogen. Hypertension 2019; 73: 1249–57.

11 Wu C-H, Wang Y, Ma M, et al. Antisense oligonucleotides targeting angiotensinogen: insights from animal studies. Biosci Rep 2019; 39. DOI:10.1042/BSR20180201.

12 A Study to Evaluate ALN-AGT01 in Patients With Hypertension. Clinicaltrials.gov. https://clinicaltrials.gov/ct2/show/NCT03934307.

13 A Study to Assess the Safety, Tolerability and Efficacy of IONIS-AGT-LRx. ClinicalTrials.gov. https://clinicaltrials.gov/ct2/show/NCT04083222.

14 A Study to Assess the Safety, Tolerability and Efficacy of IONIS-AGT-LRx, an Antisense Inhibitor Administered Subcutaneously to Hypertensive Subjects With Controlled Blood Pressure. ClinicalTrials.gov. https://clinicaltrials.gov/ct2/show/NCT03714776.

15 Desai M, Stockbridge N, Temple R. Blood pressure as an example of a biomarker that functions as a surrogate. AAPS J 2006; 8: E146-52.

16 Table of Surrogate Endpoints. U.S. Food and Drug Administration. https://www.fda.gov/drugs/development-resources/table-surrogate-endpoints-were-basis-drug-approval-or-licensure.

17 Hypertension Indication: Drug Labeling for Cardiovascular Outcome Claims. U.S. Food and Drug Administration. https://www.fda.gov/regulatory-information/search-fda-guidance-documents/hypertension-indication-drug-labeling-cardiovascular-outcome-claims.

18 Pool JL. Direct renin inhibition: focus on aliskiren. J Manag Care Pharm 2007; 13: 21–33.

19 Frampton JE, Curran MP. Aliskiren: a review of its use in the management of hypertension. Drugs 2007;67:1767-92.

20 ALLHAT Officers and Coordinators for the ALLHAT Collaborative Research Group. The Antihypertensive and Lipid-Lowering Treatment to Prevent Heart Attack Trial. Major outcomes in high-risk hypertensive patients randomized to angiotensin-converting enzyme inhibitor or calcium channel blocker vs diuretic: The Antihypertensive and Lipid-Lowering Treatment to Prevent Heart Attack Trial (ALLHAT). JAMA 2002; 288: 2981–97.

21 Dahlof B, Devereux RB, Kjeldsen SE, et al. Cardiovascular morbidity and mortality in the Losartan Intervention For Endpoint reduction in hypertension study (LIFE): a randomised trial against atenolol. Lancet 2002; 359: 995–1003.

22 Jamerson K, Weber MA, Bakris GL, et al. Benazepril plus amlodipine or hydrochlorothiazide for hypertension in high-risk patients. N Engl J Med 2008; 359: 2417–28.

23 Pantzaris N-D, Karanikolas E, Tsiotsios K, Velissaris D. Renin Inhibition with Aliskiren: A Decade of Clinical Experience. J Clin Med Res 2017; 6. DOI:10.3390/jcm6060061.

24 Holmes MV, Simon T, Exeter HJ, et al. Secretory phospholipase A(2)-IIA and cardiovascular disease: a mendelian randomization study. J Am Coll Cardiol 2013; 62: 1966–76.

25 Scott RA, Freitag DF, Li L, et al. A genomic approach to therapeutic target validation identifies a glucose-lowering GLP1R variant protective for coronary heart disease. Sci Transl Med 2016; 8: 341ra76.

26 Schmidt AF, Swerdlow DI, Holmes MV, et al. PCSK9 genetic variants and risk of type 2 diabetes: a mendelian randomisation study. Lancet Diabetes Endocrinol 2017; 5: 97–105.

27 Bovijn J, Krebs K, Chen C-Y, et al. Evaluating the cardiovascular safety of sclerostin inhibition using evidence from meta-analysis of clinical trials and human genetics. Sci Transl Med 2020; 12. DOI:10.1126/scitranslmed.aay6570.

28 Forouzanfar MH, Liu P, Roth GA, et al. Global Burden of Hypertension and Systolic Blood Pressure of at Least 110 to 115 mm Hg, 1990-2015. JAMA 2017; 317: 165–82.

29 Yang J, Ferreira T, Morris AP, et al. Conditional and joint multiple-SNP analysis of GWAS summary statistics identifies additional variants influencing complex traits. Nat Genet 2012; 44: 369–75, S1-3.

30 Gazal S, Loh P-R, Finucane HK, et al. Functional architecture of low-frequency variants highlights strength of negative selection across coding and non-coding annotations. Nat Genet 2018; 50: 1600–7.

31 Richardson TG, Davey Smith G, Munafò MR. Conditioning on a Collider May Induce Spurious Associations: Do the Results of Gale et al. (2017) Support a Health-Protective Effect of Neuroticism in Population Subgroups? Psychol. Sci. 2019; 30: 629–32.

32 Evangelou E, Warren HR, Mosen-Ansorena D, et al. Genetic analysis of over 1 million people identifies 535 new loci associated with blood pressure traits. Nat Genet 2018; 50: 1412–25.

33 Walker VM, Harrison S, Carter AR, Gill D, Tzoulaki I, Davies NM. The consequences of adjustment, correction and selection in genome-wide association studies used for two-sample Mendelian randomization. medRxiv. DOI:10.1101/2020.07.13.20152413.

34 Carvalho-Silva D, Pierleoni A, Pignatelli M, et al. Open Targets Platform: new developments and updates two years on. Nucleic Acids Res 2019; 47: D1056-65.

35 Myers TA, Chanock SJ, Machiela MJ. LDlinkR: An R Package for Rapidly Calculating Linkage Disequilibrium Statistics in Diverse Populations. Front Genet 2020; 11: 157.

36 FinnGen. Documentation of R2 release. 2020. https://finngen.gitbook.io/documentation.

37 Sudlow C, Gallacher J, Allen N, et al. UK biobank: an open access resource for identifying the causes of a wide range of complex diseases of middle and old age. PLoS Med 2015; 12: e1001779.

38 Bycroft C, Freeman C, Petkova D, et al. The UK Biobank resource with deep phenotyping and genomic data. Nature 2018; 562: 203–9.

39 Wain LV, Shrine N, Miller S, et al. Novel insights into the genetics of smoking behaviour, lung function, and chronic obstructive pulmonary disease (UK BiLEVE): a genetic association study in UK Biobank. Lancet Respir Med 2015; 3: 769–81.

40 McCarthy S, Das S, Kretzschmar W, et al. A reference panel of 64,976 haplotypes for genotype imputation. Nat Genet 2016; 48: 1279–83.

41 Tobin MD, Sheehan NA, Scurrah KJ, Burton PR. Adjusting for treatment effects in studies of quantitative traits: antihypertensive therapy and systolic blood pressure. Stat Med 2005; 24: 2911–35.

42 the CARDIoGRAMplusC4D Consortium. A comprehensive 1000 Genomes-based genome-wide association meta-analysis of coronary artery disease. Nat Genet 2015; 47: 1121.

43 Malik R, Chauhan G, Traylor M, et al. Multiancestry genome-wide association study of 520,000 subjects identifies 32 loci associated with stroke and stroke subtypes. Nat Genet 2018; 50: 524–37.

44 Woo D, Falcone GJ, Devan WJ, et al. Meta-analysis of genome-wide association studies identifies 1q22 as a susceptibility locus for intracerebral hemorrhage. Am J Hum Genet 2014; 94: 511–21.

45 Shah S, Henry A, Roselli C, et al. Genome-wide association and Mendelian randomisation analysis provide insights into the pathogenesis of heart failure. Nat Commun 2020; 11: 163.

46 Wuttke M, Li Y, Li M, et al. A catalog of genetic loci associated with kidney function from analyses of a million individuals. Nat Genet 2019; 51: 957–72.

47 Teumer A, Tin A, Sorice R, et al. Genome-wide Association Studies Identify Genetic Loci Associated With Albuminuria in Diabetes. Diabetes 2016; 65: 803–17.

48 Haas ME, Aragam KG, Emdin CA, et al. Genetic Association of Albuminuria with Cardiometabolic Disease and Blood Pressure. Am J Hum Genet 2018; 103: 461–73.

49 Willer CJ, Schmidt EM, Sengupta S, et al. Discovery and refinement of loci associated with lipid levels. Nat Genet 2013; 45: 1274–83.

50 Liu DJ, Peloso GM, Yu H, et al. Exome-wide association study of plasma lipids in >300,000 individuals. Nat Genet 2017; 49: 1758–66.

51 Giambartolomei C, Vukcevic D, Schadt EE, et al. Bayesian test for colocalisation between pairs of genetic association studies using summary statistics. PLoS Genet 2014; 10: e1004383.

52 GTEx Consortium, Laboratory, Data Analysis & Coordinating Center (LDACC)—Analysis Working Group, Statistical Methods groups—Analysis Working Group, et al. Genetic effects on gene expression across human tissues. Nature 2017; 550: 204–13.

53 Suhre K, Arnold M, Bhagwat AM, et al. Connecting genetic risk to disease end points through the human blood plasma proteome. Nat Commun 2017; 8: 14357.

54 Alasoo K, Rodrigues J, Mukhopadhyay S, et al. Shared genetic effects on chromatin and gene expression indicate a role for enhancer priming in immune response. Nat Genet 2018; 50: 424–31.

55 Çalişkan M, Manduchi E, Rao HS, et al. Genetic and Epigenetic Fine Mapping of Complex Trait Associated Loci in the Human Liver. Am J Hum Genet 2019; 105: 89–107.

56 Siewert KM, Voight BF. Bivariate Genome-Wide Association Scan Identifies 6 Novel Loci Associated With Lipid Levels and Coronary Artery Disease. Circ Genom Precis Med 2018; 11: e002239.

57 Huckins LM, Dobbyn A, Ruderfer DM, et al. Gene expression imputation across multiple brain regions provides insights into schizophrenia risk. Nat Genet 2019; 51: 659–74.

58 Sabik OL, Calabrese GM, Taleghani E, Ackert-Bicknell CL, Farber CR. Identification of a core module for bone mineral density through the integration of a co-expression network and GWAS data. bioRxiv. DOI:10.1101/803197.

59 Andaleon A, Mogil LS, Wheeler HE. Genetically regulated gene expression underlies lipid traits in Hispanic cohorts. PLoS One 2019; 14: e0220827.

60 Liu B, Gloudemans MJ, Rao AS, Ingelsson E, Montgomery SB. Abundant associations with gene expression complicate GWAS follow-up. Nat Genet 2019; 51: 768–9.

61 Viechtbauer W. Conducting meta-analyses in R with the metafor package. J Stat Softw 2010; 36. DOI:10.18637/jss.v036.i03.

62 Walker VM, Davies NM, Hemani G, et al. Using the MR-Base platform to investigate risk factors and drug targets for thousands of phenotypes. Wellcome Open Res 2019; 4: 113.

63 Yavorska OO, Burgess S. MendelianRandomization: an R package for performing Mendelian randomization analyses using summarized data. Int J Epidemiol 2017; 46: 1734–9.

64 Zhou W, Nielsen JB, Fritsche LG, et al. Efficiently controlling for case-control imbalance and sample relatedness in large-scale genetic association studies. Nat Genet 2018; 50: 1335–41.

65 UK Biobank — Neale lab. Neale lab. http://www.nealelab.is/uk-biobank.

66 Ettehad D, Emdin CA, Kiran A, et al. Blood pressure lowering for prevention of cardiovascular disease and death: a systematic review and meta-analysis. Lancet 2016; 387: 957–67.

67 Wright JM, Musini VM, Gill R. First- line drugs for hypertension. Cochrane Database Syst Rev 2018. DOI:10.1002/14651858.CD001841.pub3.

68 Alnylam Pharmaceuticals Reports First Quarter 2020 Financial Results and Highlights Recent Period Activity. Alnylam Pharmaceuticals, Inc. https://investors.alnylam.com/press-release?id=24811.

69 Ference BA. How to use Mendelian randomization to anticipate the results of randomized trials. Eur Heart J 2018; 39: 360–2.

70 Hackam DG, Redelmeier DA. Translation of research evidence from animals to humans. JAMA 2006; 296: 1731–2.

71 Perel P, Roberts I, Sena E, et al. Comparison of treatment effects between animal experiments and clinical trials: systematic review. BMJ; 334: 197.

72 Davies NM, Holmes MV, Davey Smith G. Reading Mendelian randomisation studies: a guide, glossary, and checklist for clinicians. BMJ 2018; 362: k601.

73 Holmes MV. Human Genetics and Drug Development. N Engl J Med 2019; 380: 1076–9.

74 Georgakis Marios K., Malik Rainer, Gill Dipender, et al. Interleukin-6 Signaling Effects on Ischemic Stroke and Other Cardiovascular Outcomes. Circulation: Genomic and Precision Medicine 2020; 13: e002872.

75 Bick AG, Pirruccello JP, Griffin GK, et al. Genetic Interleukin 6 Signaling Deficiency Attenuates Cardiovascular Risk in Clonal Hematopoiesis. Circulation 2020; 141: 124–31.

76 Myocardial Infarction Genetics Consortium Investigators, Stitziel NO, Won H-H, et al. Inactivating mutations in NPC1L1 and protection from coronary heart disease. N Engl J Med 2014; 371: 2072–82.

77 Gill D, Georgakis MK, Koskeridis F, et al. Use of Genetic Variants Related to Antihypertensive Drugs to Inform on Efficacy and Side Effects. Circulation 2019; 140: 270–9.

78 Kunz R, Friedrich C, Wolbers M, Mann JFE. Meta-analysis: effect of monotherapy and combination therapy with inhibitors of the renin angiotensin system on proteinuria in renal disease. Ann Intern Med 2008; 148: 30–48.

79 Dickson Matthew E., Sigmund Curt D. Genetic Basis of Hypertension. Hypertension 2006; 48: 14–20.

80 Duncan LE, Ostacher M, Ballon J. How genome-wide association studies (GWAS) made traditional candidate gene studies obsolete. Neuropsychopharmacology 2019; 44: 1518–23.

81 Watts JK, Corey DR. Silencing disease genes in the laboratory and the clinic. J Pathol 2012; 226: 365–79.

82 Bovijn J, Censin JC, Lindgren CM, Holmes MV. Using human genetics to guide the repurposing of medicines. Int J Epidemiol 2020; published online Feb 25. DOI:10.1093/ije/dyaa015.

83 Millwood IY, Bennett DA, Holmes MV, et al. Association of CETP Gene Variants With Risk for Vascular and Nonvascular Diseases Among Chinese Adults. JAMA Cardiol 2017; 3: 34–43.

84 Schmidt AF, Holmes MV, Preiss D, et al. Phenome-wide association analysis of LDL-cholesterol lowering genetic variants in PCSK9. BMC Cardiovasc Disord 2019; 19: 240.

85 Sun BB, Maranville JC, Peters JE, et al. Genomic atlas of the human plasma proteome. Nature 2018; 558: 73–9.

86 Joshi A, Mayr M. In Aptamers They Trust: The Caveats of the SOMAscan Biomarker Discovery Platform from SomaLogic. Circulation 2018; 138: 2482–5.

87 Bouwens E, van den Berg VJ, Akkerhuis KM, et al. Circulating Biomarkers of Cell Adhesion Predict Clinical Outcome in Patients with Chronic Heart Failure. J Clin Med Res 2020; 9. DOI:10.3390/jcm9010195.

88 Lu H, Wu C, Howatt DA, et al. Angiotensinogen Exerts Effects Independent of Angiotensin II. Arterioscler Thromb Vasc Biol 2016; 36: 256–65.

89 Schmidt AF, Finan C, Gordillo-Marañón M, et al. Genetic drug target validation using Mendelian randomisation. Nat Commun 2020; 11: 3255.

90 Richardson TG, Sanderson E, Palmer TM, et al. Evaluating the relationship between circulating lipoprotein lipids and apolipoproteins with risk of coronary heart disease: A multivariable Mendelian randomisation analysis. PLoS Med 2020; 17: e1003062.

91 Armitage J, Holmes MV, Preiss D. Cholesteryl Ester Transfer Protein Inhibition for Preventing Cardiovascular Events: JACC Review Topic of the Week. J Am Coll Cardiol 2019; 73: 477–87.

92 Khetarpal SA, Wang M, Khera AV. Volanesorsen, Familial Chylomicronemia Syndrome, and Thrombocytopenia. N. Engl. J. Med. 2019; 381: 2584.

93 Witztum JL, Gaudet D, Freedman SD, et al. Volanesorsen and Triglyceride Levels in Familial Chylomicronemia Syndrome. N Engl J Med 2019; 381: 531–42.

94 Akcea and Ionis report positive topline Phase 2 results of AKCEA-APOCIII-L Rx | Akcea Therapeutics. Akcea Therapeutics. https://ir.akceatx.com/news-releases/news-release-details/akcea-and-ionis-report-positive-topline-phase-2-results-akcea.

95 Mazzaglia G, Ambrosioni E, Alacqua M, et al. Adherence to antihypertensive medications and cardiovascular morbidity among newly diagnosed hypertensive patients. Circulation 2009; 120: 1598–605.

